# Inherited extremes of aortic diameter confer risk for a specific class of congenital heart disease

**DOI:** 10.1101/2020.08.12.20172817

**Authors:** Catherine Tcheandjieu, Daniela Zanetti, Mengyao Yu, James R. Priest

## Abstract

Population based studies demonstrate strong familial recurrence of cardiac malformations particularly for individuals affected with a specific class of CHD, left-ventricular outflow tract (LVOT) obstruction. Recently we linked 99 lead variants across 71 loci to diameter of the ascending aorta derived from MRI measurements across multiple ethnicities in the UK Biobank. Using these data we created a polygenic risk score capturing ascending aortic diameter (PRSAoD) Among 2,594 individuals with CHD; a decrease by one standard deviation in PRSAoD was associated with an increased risk of congenital LVOT (OR=1.14[1.03-1.26], p=0.014) but not with other subtypes of CHD. Using Mendelian Randomization we observed strong evidence of a causal effect where inheritance of a smaller diameter of the ascending aorta corresponded to an increase in risk for congenital LVOT (p_IVW = 0.008). Our data may suggest that genetic determinants of a smaller ascending aorta act during early development to disturb blood flow through left-sided structures to increasing the risk of LVOT CHD, which is consistent with experimental evidence and the “no flow, no grow” paradigm in the formation of the left ventricular outflow tract.

## Manuscript

Congenital heart disease (CHD) is the most common anatomical malformation and while the risk of CHD is highly heritable, *de novo* mutations or inheritance of pathogenic coding variation across all known genes related to CHD identify less than 10% of the risk for disease. Therefore, important genetic risk factors for CHD remain to be discovered.

Population based studies demonstrate strong familial recurrence of cardiac malformations with a relative risk of 12.9 for first-degree family members of individuals affected with a specific class of CHD, left-ventricular outflow tract (LVOT) obstruction^1^. Within families, highly heritable differences in the size of the aortic root (polygenic heritability-0.96)^2^ have been observed amongst unaffected first-degree family members of individuals with LVOT CHD. Experimental restriction of ascending aortic diameter in fetal lambs has been shown to be sufficient to cause severe forms of LVOT CHD^3^. These clinical and experimental observations suggest that heritable extremes of normal variation in cardiac anatomy may be related to rare forms of CHD such as LVOT.

Recently we linked 99 lead variants across 71 loci to diameter of the ascending aorta derived from MRI measurements across multiple ethnicities in the UK Biobank^4^. Using these data we created a polygenic risk score capturing ascending aortic diameter (PRS_AoD_) and applied it to a British study of individual level data for 2,594 individuals with five different forms of CHD and 5,159 adult controls drawn from the Welcome Trust Case Control Consortium^5^. Among the 2,594 individuals with CHD; a decrease by one standard deviation in PRS_AoD_ was associated with an increased risk of congenital LVOT (OR=1.14[1.03-1.26], p=0.014) but not with other subtypes of CHD [Fig.1A]. This observation suggests that relative to other subtypes of CHD, individuals affected with LVOT may have inherited a smaller genetically determined size of the ascending aorta than would be expected by chance.

**Figure 1.**
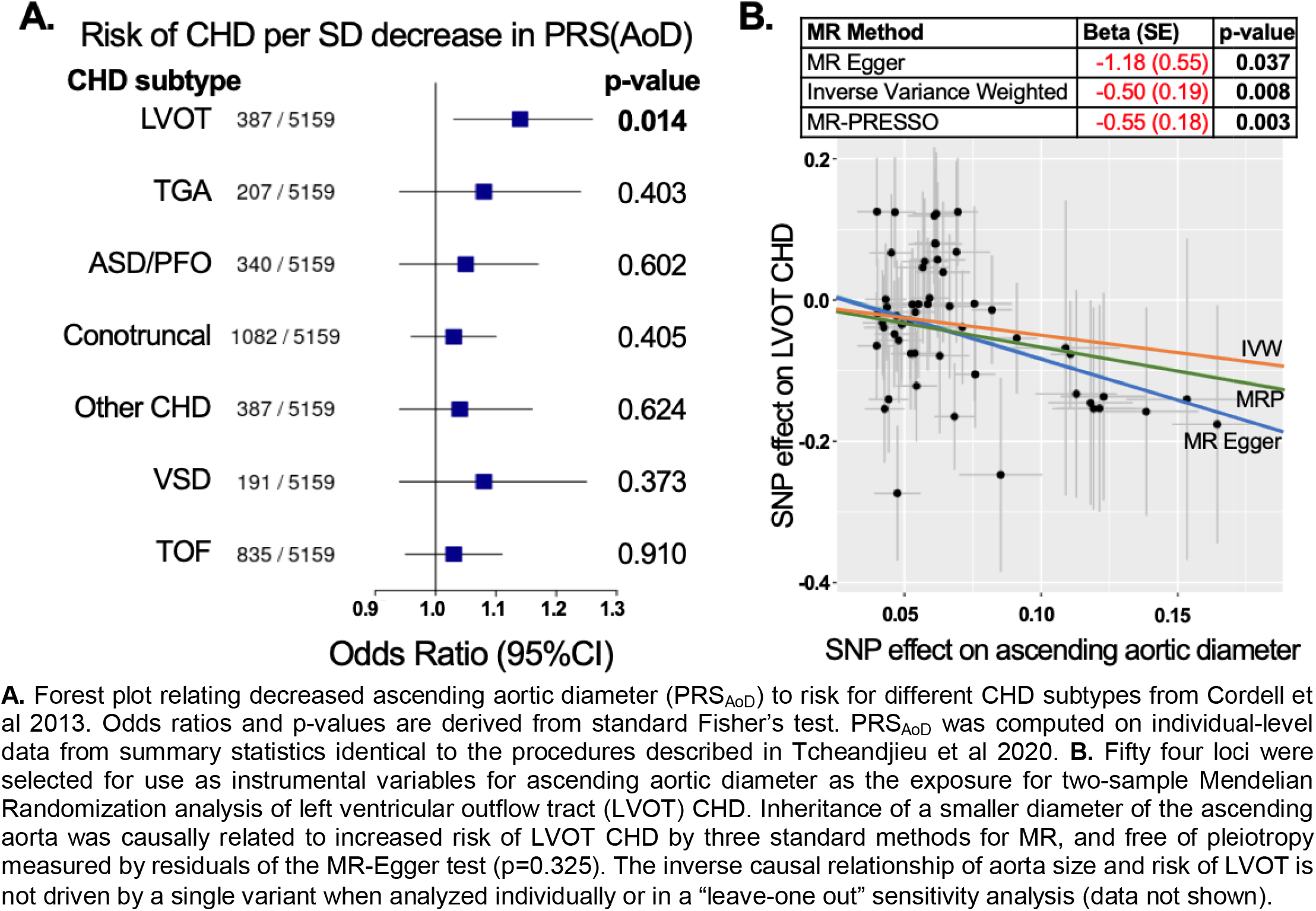
Inheritance of a smaller ascending aorta is a causal risk factor for LVOT CHD.A.

To specifically test for a causal relationship between inheriting the genetic liability for a small ascending aortic diameter and increased risk of LVOT suggested by experimental studies^3^, we performed two-sample Mendelian randomization (MR) as implemented in R language for statistical computing (https://mrcieu.github.io/TwoSampleMR/) using separate GWAS for exposure and outcome. Taking the 99 variants with p < 5e-08 from a trans-ethnic meta-analysis of ascending aortic diameter^4^, we performed clumping using r^2^ of 0.001 across a window of 100 kb to create an instrumental variable (IV) for use in MR. To avoid horizontal pleiotropy we excluded variants directly related to LVOT with p < 0.01. For the outcome GWAS for LVOT, we performed re-imputation of 387 individuals with LVOT and 5159 controls to the TOPMed reference panel to obtain the 54 variants included in the IV and performed standard logistic regression including genetic sex as a covariate. We estimated statistical power to detect a causal relationship was better than 80% using variance explained from our exposure IV, effect size from observational analyses^1^, and an α of 0.05 using a standard online calculator (https://sb452.shinyapps.io/power/).

Across three MR methods tested, we observed strong evidence of a causal effect where inheritance of a smaller diameter of the ascending aorta corresponded to an increase in risk for LVOT **[Fig.1B]**. Tests for heterogeneity did not suggest important differences in effect size or direction (p_Q-IVW_=0.67) while sensitivity analyses suggested the observed causal effect was relatively uniform across the 54 variants included in the instrumental variable, and there was no evidence of horizontal pleiotropy by a global test for outliers (p=0.68) (https://github.com/rondolab/MR-PRESSO).

Taken together, our findings suggest that common inherited genetic determinants of smaller ascending aortic diameter increase the risk of congenital LVOT. This observation is consistent with experimental models of cardiac development and supports the paradigm of “no flow, no grow” in the formation and growth of the left ventricular outflow tract^3^. Our data may suggest that genetic determinants of a smaller ascending aorta act during early development to disturb blood flow through left-sided structures thereby increasing the risk of LVOT CHD. More broadly, these data may represent a newly recognized heritable form of risk for CHD that originates from inherited extremes in the size of developing cardiovascular anatomy. Within a population where the size of anatomical structures displays a Normal distribution, such extremes in phenotype will arise from the random inheritance of common variation as demonstrated for other anatomical traits and cardiovascular diseases^4^. Overall these data may provide evidence supporting a model in which inherited extremes in the size of anatomical structures in the heart and large vessels represent causal factors for rare forms of CHD.

## Data Availability

Data from the ascending aorta is available online while data on individuals with congenital heart disease is available upon request from Dr. Heather Cordell.

## Acknowledgments

This research has been conducted using the UK Biobank resource under application number 15860.

## REFERENCES

1. Øyen N, Poulsen G, Boyd HA, Wohlfahrt J, Jensen PKA, Melbye M. Recurrence of congenital heart defects in families. Circulation [Internet]. 2009;120:295-301. Available from: http://circ.ahajournals.org/content/120/47295.long

2. McBride KL, Pignatelli R, Lewin M, Ho T, Fernbach S, Menesses A, Lam W, Leal SM, Kaplan N, Schliekelman P, Towbin JA, Belmont JW. Inheritance analysis of congenital left ventricular outflow tract obstruction malformations: Segregation, multiplex relative risk, and heritability. Am J Med Genet Part A [Internet]. 2005 [cited 2019 Jul 17];134A:180–186. Available from: http://www.ncbi.nlm.nih.gov/pubmed/15690347

3. Fishman NH, Hof RB, Rudolph AM, Heymann MA. Models of congenital heart disease in fetal lambs. Circulation [Internet]. 1978 [cited 2020 Jul 21];58:354–364. Available from: https://www.ahajournals.org/doi/10.1161/01.CIR.58.2.354

4. Tcheandjieu C, Xiao K, Tejeda H, Lynch J, Ruotsalainen S, Bellomo T, Palnati M, Judy R, Kember R, Klarin D, Kember R, Verma S, Center RG, Program VMV, Project F, Palotie A, Daly M, Ritchie M, Rader D, Rivas MA, Assimes T, Tsao P, Damrauer S, Priest J. High heritability of ascending aortic diameter and multi-ethnic prediction of thoracic aortic disease. *medRxiv* [Internet]. 2020 [cited 2020 Jul 21];14:2020.05.29.20102335. Available from: https://www.medrxiv.org/content/10.1101/2020.05.29.20102335v1

5. Cordell HJ, Bentham J, Topf A, Zelenika D, Heath S, Mamasoula C, Cosgrove C, Blue G, Granados-Riveron J, Setchfield K, Thornborough C, Breckpot J, Soemedi R, Martin R, Rahman TJ, Hall D, van Engelen K, Moorman AFM, Zwinderman AH, Barnett P, Koopmann TT, Adriaens ME, Varro A, George AL, dos Remedios C, Bishopric NH, Bezzina CR, O’Sullivan J, Gewillig M, Bu’Lock FA, Winlaw D, Bhattacharya S, Devriendt K, Brook JD, Mulder BJM, Mital S, Postma A V, Lathrop GM, Farrall M, Goodship JA, Keavney BD, O’Sullivan J, Gewillig M, Bu’Lock FA, Winlaw D, Bhattacharya S, Devriendt K, Brook JD, Mulder BJM, Mital S, Postma A V, Lathrop GM, Farrall M, Goodship JA, Keavney BD. Genome-wide association study of multiple congenital heart disease phenotypes identifies a susceptibility locus for atrial septal defect at chromosome 4p16. Nat Genet [Internet]. 2013;45:822–4. Available from: http://www.pubmedcentral.nih.gov/articlerender.fcgi?artid=3793630&tool=pmcentrez&rendertype=abstract

